# Support and follow-up needs of patients discharged from Intensive Care after severe COVID-19: a mixed-methods study of the views of UK general practitioners and intensive care staff during the pandemic’s first wave

**DOI:** 10.1101/2020.12.23.20248798

**Authors:** Ana Castro-Avila, Laura Jefferson, Veronica Dale, Karen Bloor

## Abstract

**Background:** After discharge from an intensive care unit(ICU), over 50% of patients face significant physical, mental and cognitive problems. The COVID-19 pandemic has resulted in a large cohort of these patients who will need follow-up services that can address their support needs.

**Objectives:** To identify follow-up services planned for COVID-19 patients discharged from ICU, and to explore the views of ICU staff and General Practitioners(GPs) regarding these patients’ future needs and care coordination.

**Design:** Sequential mixed-methods UK study. We explored usual follow-up practice after ICU discharge and changes in follow-up during the pandemic through a survey of ICU staff, and GP awareness of follow-up and support needs of patients discharged from ICU through a survey of GPs. Through these surveys, we identified participants for semi-structured interviews with both groups.

**Results:** We obtained 170 survey responses and conducted 23 interviews. Over 60% of GPs were unaware of the follow-up services generally provided by their local hospitals, and whether or not these were functioning during the pandemic. Eighty percent of ICUs reported some form of follow-up services, with 25% of these suspending provision during the peak of the pandemic, and over half modifying their provision (usually to provide the service remotely). Complex funding streams, poor communication between primary and secondary care, and lack of clarity about who was responsible for referrals and follow-up were the most common problems identified. Discharge documentation was described as poor and lacking key information. Both groups mentioned difficulties accessing services in the community.

**Conclusions:** The pandemic has highlighted long-standing issues of continuity of care and complex funding streams for post-ICU follow-up care. The large cohort of ICU patients admitted due to COVID-19 highlights the need for improved follow-up services and communication between specialists and GPs, not only for COVID-19 patients, but for all those discharged from ICU.

**Article Summary:** *Strengths and limitations of this study:*

- This is the first study exploring NHS staff views of post-ICU follow-up services to support patients recovering from severe COVID-19.
- Responses to our survey had good geographic spread but were limited in number and relied on volunteers.
- GP interviews were small in number, but our findings align with those of larger studies conducted before the pandemic.

## Introduction

The significant physical, mental and cognitive problems patients face following a stay in an Intensive Care Unit (ICU) are well-documented.^1 2^ Including prolonged muscle weakness, cognitive dysfunction and symptoms of post-traumatic stress disorder (PTSD), these are often collectively referred to as ‘post intensive care syndrome’ (PICS)^3^ and may persist even five years after leaving hospital.^4-7^ Six months post-discharge, 25% of survivors suffer severe disability,^8^ and only around 55% have returned to work.^9^ Psychological disorders including depression, anxiety, and post-traumatic stress disorder (PTSD) are common, affecting 55% in the first year following ICU discharge.^10^ The variety and severity of sequelae vary substantially.

The COVID-19 pandemic has resulted in a large and rapid increase in intensive care activity, which will challenge post-ICU services in several ways. Increases in ICU capacity necessitated the use of less experienced staff and very high workloads. More stringent infection control protocols created new potential causes of distress, including not allowing family members inside the unit and health care professionals wearing Personal Protective Equipment (PPE). These factors might lead to a very large cohort of critical illness survivors that might have greater than expected needs due to the context and length of their critical care stay, ^11^ which could put the current capacity of services under stress.

Post-ICU follow-up from hospital teams is likely to have been compromised during the pandemic. The availability, form and scale of services for ICU survivors of COVID-19 are largely unknown, and patient needs are difficult to predict. Timely, appropriate support could potentially prevent future problems in patients’ physical, cognitive and mental health and care. This study aims to identify follow-up services that were available during and after the UK’s first wave, early reflections on care during the first wave and the views of critical care staff and GPs regarding the future needs of COVID-19 patients.

This study aims to identify follow-up services planned for COVID-19 patients discharged from ICU, and to explore the views of ICU staff and General Practitioners (GPs) regarding these patients’ future needs and care coordination.

## Methods

We employed mixed methods including online surveys and semi-structured telephone interviews with ICU follow-up lead clinicians and general practitioners (GPs).

In collaboration with clinicians in the field, we developed a very brief questionnaire of ICU staff (Table 1) to determine usual procedures of follow-up of patients after ICU discharge, and changes during the pandemic. The Intensive Care Society, Faculty of Intensive Care Medicine, British Association of Critical Care Nurses, and leading experts on intensive care disseminated this survey through newsletters, targeted emails and Twitter. We invited clinicians managing follow-up care of discharged ICU patients to participate, and asked respondents to volunteer for interviews. We sampled volunteers purposively by geographical location, need for expanded bed capacity during the pandemic, and changes implemented.

**Table 1.**
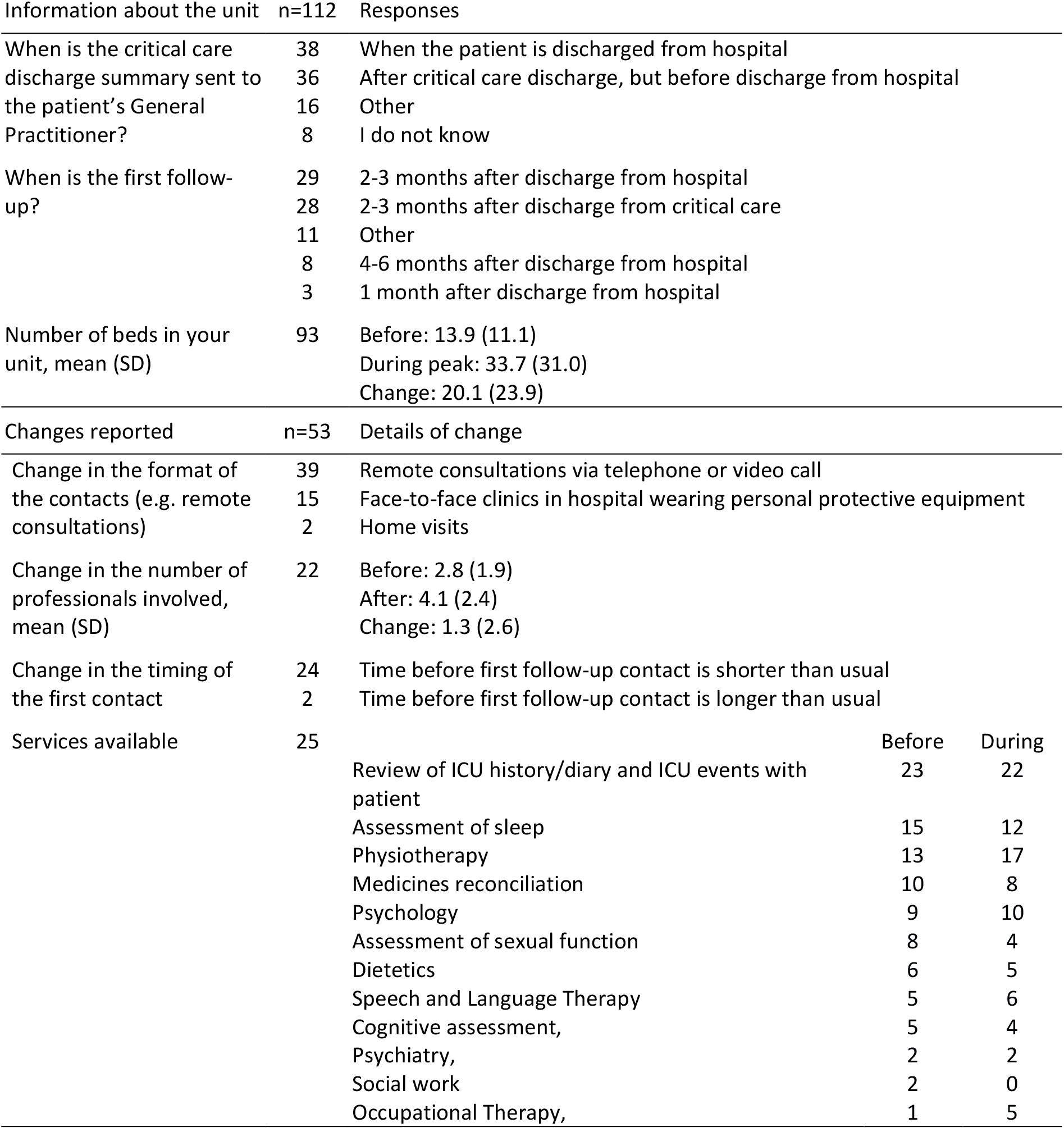
Responses of ICU leads about follow-up services during the pandemic

In collaboration with the Royal College of General Practitioners (RCGP), we developed and distributed a very brief questionnaire (Table 2) exploring GPs’ awareness of post-ICU follow-up services, and broad concerns about severe COVID-19 patients’ care. RCGP also included three of these questions in a routine survey of their GP research panel. GPs proved difficult to recruit to interviews through the survey, so we supplemented this with ‘snowballing’ using contacts at the RCGP, University of York and The King’s Fund. We attempted to generate geographical spread in terms of location and COVID-19 incidence.

**Table 2.** Responses from GPs survey

| Question | Responses | Our sample | RCGP |
| --- | --- | --- | --- |
| Does your nearest Hospital Trust have specific follow-up services for all patients who have been discharged from critical care? | n | 58 | - |
|  | I do not know | 36 (62%) | - |
|  | Yes | 11 (19%) | - |
|  | No | 11 (19%) | - |
| Is the follow-up service functioning during the COVID-19 pandemic? | n | 45 | - |
|  | I do not know | 39 (87%) | - |
|  | Yes | 6 (13%) | - |
| Within your patient list, are you aware of any patients who have required critical care for severe COVID-19? | n | 56 | 537 |
|  | Yes | 33 (59%) | 208 (39%) |
|  | No | 13 (23%) | 244 (45%) |
|  | I do not know | 10 (18%) | 85 (16%) |
| How many of your patients went through critical care due to severe COVID-19? | n | 24 | 462 |
|  | Mean (min-max) | 4.4 (1-20) | 4.4 (0-50) |
| Considering future patients in your practice recovering from a COVID-19 related critical care stay, please rank your concerns about their care, mean rank (SD) | n | 40 | 447 |
|  | Physical health care | 1.9 (1.4) | 1.4 (1.3) |
|  | Mental Health care | 2.4 (1.1) | 1.4 (1.2) |
|  | Access to rehabilitation services | 3.1 (1.3) | 1.6 (1.3) |
|  | Cognitive functioning | 3.5 (1.2) | 1.8 (1.3) |
|  | Access to social care | 4.0 (1.1) | 1.7 (1.4) |

Both surveys were piloted with clinicians and experts to ensure clarity, conciseness, precision of language, and identify any essential omissions. We used Qualtrics XM software (Qualtrics, Provo, UT) to distribute the surveys and collect responses.

We conducted 30-minute semi-structured telephone interviews with GPs and ICU leads, which were audio-recorded for analysis. We asked for consent verbally before the start of each interview. Interviews with ICU leads explored views on whether and how the future needs of COVID-19 patients differed from non-COVID patients and captured early reflections on ICU care and transitions during the first wave of the COVID-19 outbreak. Interviews with GPs explored their prior experience of managing post-ICU patients, and information needs in relation to severe COVID-19.

### Data analysis

#### Quantitative

Survey data were exported to IBM SPSS Statistics for Windows (Version 26. Armonk, NY: IBM Corp) for analysis. Absolute and relative frequencies were used to summarise responses. We calculated the average rank for the question about GPs’ concerns regarding future care needs of patients recovering from a COVID-19 related critical care stay.

#### Qualitative

We conducted thematic analysis to synthesise the results and produce insights using NVivo (version 12. QSR International Pty Ltd.) As is increasingly being adopted in rapid qualitative research,^12^ analysis was undertaken directly from audio-recordings and detailed notes, transcribing sections for use as quotations. Two researchers (ACA and LJ) coded, each producing an initial framework with main themes, which were discussed with the wider team and topic experts to refine the framework and distil overarching themes. Some representative quotes are presented to contextualise and aid interpretation.

### Patient and Public Involvement (PPI)

We reviewed and discussed the project protocol with our PPI group, refining it in response. We were, however, unable to capture patients’ views on follow-up services in our study timescale.

## Results

### Surveys

Between 15^th^ June and 3^rd^ August, ICU follow-up lead clinicians from 112 units (43% of Acute NHS Trusts in England) responded to our survey. 83% were based in England, and 96% were from mixed intensive care and high dependency units. On average, units more than doubled bed capacity at the height of the first wave (Table 1).

Follow-up services were offered in 80 units (71% of those sampled); of those, 20 reported ceasing provision and 53 modifying provision of services during the pandemic. Eight units implemented a new follow-up service after the peak of the pandemic. Provision of occupational therapy and physiotherapy were the services with the greatest increase (Table 1).

58 GPs responded to our survey and an additional 537 responded to three questions distributed by RCGP (Table 2). 78% of RCGP responses came from England, 61% were female, 63% were 35-54 years old, and 83% were white.

### Interview findings

We conducted 23 interviews between 23^rd^ June and 30^th^ July; 17 with ICU staff (7 ICU consultants, 7 senior nurses, 3 rehabilitation coordinators) and 6 GPs. ICU interviews covered all UK regions, with the ICUs having an average capacity before the pandemic of 14 beds (range 4-60); increasing by 16 beds on average (range 2-38 beds). The GPs covered different regions of England and a mix of patient demographics.

### The ICU environment

All interviewees reported opening new areas and bringing nurses from other areas (e.g. theatre, surgical recovery, other hospital wards) particularly those with ICU training. Consultants increased the frequency of their rotations to ensure continuous coverage. Administrative tasks for clinicians were suspended and all staff providing outpatient or outreach services returned to inpatient activities. ICU nurses split their time between patient care, staff supervision and training new staff, which was reported to increase workload and stress. Hospitals with greater bed capacity implemented proning and intubating teams, and some implemented retrieval teams to transfer patients between hospitals.

ICU staff reported that PPE was uncomfortable and created difficulties in building relationships with patients, hindering instructions and explanations. Visiting of patients’ family members was suspended, except at end of life; therefore, staff implemented different strategies to update families and enable virtual visiting.

Numerous ICU interviewees mentioned that patients with COVID-19 may represent a new patient group, but are still ICU survivors, with the weakness, mental and cognitive problems these patients commonly suffer. They expected COVID-19 patients to suffer a longer-lasting deterioration of lung function, potential issues with renal function, a high incidence of shoulder injuries due to proning, and cognitive problems related to the incidence of delirium.

Some thought it was too early to tell whether they will experience more physiological and psychological problems, but many highlighted particular treatments, including prolonged and deep sedation, opioids and neuromuscular blockers, which are associated with increased risk of muscular weakness, polyneuropathy, and cognitive impairments. Patients experienced extended periods in a prone position, mechanical ventilation and less experienced nursing staff. One consultant believed that actively screening for mental health problems was needed (ICUcons09, Scotland).

One ICU nurse had administered a mental health questionnaire with COVID-19 patients as part of patient audit, reporting that ventilated patients had the same psychological issues as other ICU patients, but those who received continuous positive airways pressure (CPAP) and were therefore conscious, had worse scores. An ICU consultant echoed this and also highlighted potential difficulties due to PPE:

> *“The other people in the bay watched [another patient] die over a number of days… It doesn’t surprise me that the people here perhaps, more awake and aware are very, very traumatised by the experiences […] [Some patients] have delusional thoughts. I mean, I think that’s gonna be a lot worse when you’re surrounded by someone wearing a hazmat suit*.*” ICUcons06, South West*.

### Provision of follow-up services

Before COVID-19, most ICU interviewees reported having a post-ICU follow-up service; the few that did not were planning to implement one after the pandemic. Most follow-up services were suspended during the peak of the first wave, as staff returned to in-hospital clinical duties. The few places that continued to provide such services used telephone follow-up, delivered by staff that were shielding.

Reported provision varies greatly, with some units delivering follow-up with just a consultant and/or a senior nurse, while others have multidisciplinary teams. Some units start their follow-up during the ICU stay, and have designated professionals to assess, refer and follow patients during the hospital episode and into the community. Others with well-established follow-up services refer ICU patients to pulmonary or cardiac rehab to recover fitness and muscle strength.

All unit staff we interviewed follow patients up 2-3 months after ICU discharge, but a minority also routinely call patients weekly (ICUnurse04, North East) or monthly (ICUnurse08, East Midlands). All had to change the format of their follow-up during the pandemic, most replaced clinics with telephone calls or virtual consultations. One senior nurse highlighted the challenges of reduced non-verbal communication and time-limited calls:

> *“Phone calls don’t really cut it because unless you’re very skilled at talking to people, assessing people, you’re not going to pick up on all those cues that people give out […] if we’ve got half an hour appointment, we won’t get much out from in 10 minutes, but they’ll open up” ICUnurse14, East of England*

Two ICU interviewees said that they were implementing separate clinics for COVID-19 patients to carry out extra recommended assessments, such as a chest x-ray at 6 weeks post-discharge as recommended by the British Thoracic Society.^13^

In some locations COVID-19 rehab hospitals have been set up to provide specialist care and a “step down” for patients *“that are not quite well enough to leave the acute setting and not quite well enough to go home”* (GP1003, Yorkshire). This provided the opportunity for expert care to be delivered but relied on CCG funding and *“proactive planning for the worst-case scenario”* (GP1003, Yorkshire).

GPs were concerned about the complex psychological needs of patients recovering from severe COVID and that greater emphasis is placed on the physical needs of patients, with insufficient consideration of psychological support. All ICU interviewees agreed about the need for increased psychological support services.

Some ICU interviewees questioned the capacity of community rehabilitation services to improve patients’ functioning. Both GPs and ICU staff felt that previous notions of thresholds for functional status post-ICU (several commented on assessments of patients climbing a flight of stairs) were arbitrary and not suitable for the wider age group of patients affected by COVID-19.

> *“Community Services work at getting someone functional. They don’t work at getting them back to the state that they were at before they came into hospital. So, considering that a lot of our patients were younger patients, walking with a Zimmer frame to and from a bathroom aren’t really what they want to be doing. They want to be getting back to their fitness level and back to work*.*” ICUrehab15, Wales*

One GP commented that: *“it’s only once they are home that the true level of need is understood”* (GP1005, North West); at this point primary care and community services need to step in, but the support needs of these patients may be beyond their expertise and the capacity of services.

Some GP practices had taken proactive steps to follow-up patients discharged from ICU (before and since COVID-19). One practice, pre-COVID-19, developed a list of ‘at risk’ patients that was monitored by a nurse practitioner and discussed at daily practice meetings, with the aim of reducing hospital admissions. One London practice had instigated weekly follow-up calls with COVID-19 patients discharged from ICU, following a ‘near miss’ event whereby a serious complication had been detected opportunistically during a GP follow-up call.

### Barriers to provision of follow-up services

Both ICU staff and GPs found referrals to community follow-up services difficult, with differences in opinion about whose responsibility this was, as well as problems with waiting lists (particularly for mental health services). Community rehabilitation services were described as *“patchy”* (ICUnurse08, East Midlands). Key barriers expressed relate to funding complexities, remit and expertise, and communication.

ICU interviewees in England felt the lack of a tariff for funding ICU follow-up clinics created variation in service provision. Both ICU and GP interviewees believed that community teams similar to those for stroke or cardiac rehabilitation should be set up for post-ICU patients.

Several interviewees were concerned that already overstretched community services with existing waiting lists could not meet the increase in demand from COVID-19 without improved funding and infrastructure. One GP described the closure of some community services, and ICU interviewees had concerns that those discharged from ICU in the peak of the pandemic did not have anywhere to go.

> *“We’ve had patients in tears, we’ve had seven patients through telephone calls. And all of them are absolutely distraught and feel like they’ve been abandoned in the community […] because they were thrown out of hospital very quickly. There’s no services in the community for them at all*.*” ICUrehab15, Wales*

Both ICU and GP interviewees felt that hospital services were better placed to follow-up patients discharged from ICU, because they have a better understanding of the patient’s needs. There appears, however, to be a general lack of awareness about the difficulties of coordinating patients’ needs in each setting.

> *“I’m not sure the hospitals are always very aware of what services are available in the community… To give you a COVID example, I had a doctor ring me up and say ‘he’s been in hospital for a long time, can you make sure he sees a psychiatrist when he comes out?’… ‘no, I don’t have that kind of access to psychiatrists’*.*” GP1001, East*

Poor or delayed communication can result in misunderstandings about patients’ support needs, and GP interviews highlighted that these high-risk patients could potentially suffer adverse events if their follow-up is not adequate. Hospitals and GPs communicate through discharge letters, which all GPs described as inadequate, often produced by junior doctors and lacking pertinent detail - “*the nuance, the detail is often missing*” (GP1005, North West).

Interviewees also reported examples of good practice, for example respiratory consultants sharing contact details and working closely with GPs when severe COVID-19 patients are discharged. In some cases, ICU interviewees commented on the importance of long-standing professional relationships with community rehabilitation service providers.

GPs were concerned about the evolving nature of COVID-19 and changing medical understanding. They welcomed specific and targeted information that would help them to guide patient’s care after an intensive care and hospital stay. Others suggested a need for better communication with hospital teams to develop their understanding of specific patients’ needs, and where to find support. One GP summarised the information needs as *“what to look for, when to refer back into hospital and types of patients that need specific follow-up” (GP1002, Yorkshire)*.

All GPs stressed that guidance needs be balanced and channelled through a respected national body, as they faced an overload of information, particularly during the early phases of the pandemic when sometimes conflicting information was disseminated daily, from multiple sources.

To cope with the levels of information during the first wave, GP practices had initiated daily team meetings to discuss and keep abreast of key changes. One GP commented that the vast amount of COVID-19 information hampered GPs from employing their ‘generalist skills’ to tailor care to the individual’s needs (GP1005, North West).

### The pandemic as an opportunity to change

Interviewees from three ICUs described the pandemic as an opportunity to initiate a multidisciplinary team (MDT) follow-up clinic, by making visible the issues faced by patients discharged from ICU.

> *“[We’ve had an] uplift in the therapy staff […][as] we’ve now got more dietician[s], more physio, more pharmacy, we’ve never had an OT before six months ago, we’ve never had a psychologist of our own […] We’re now in a position to offer MDT follow-up service rather than just a simple follow-up clinic*.*” ICUcons06, South West*.

Interviewees highlighted the need for increased provision in response to the pandemic, resulting from large numbers of newly affected patients, uncertainties in their support needs, and a younger population needing to return to work.

## Discussion

### Statement of principal findings

The peak of the first wave of COVID-19 saw dramatic changes in ICUs to increase bed capacity. This was accompanied by adaptations to (and, in general, reductions in) the follow-up care provided, although most units retained some form of follow-up service.

Before COVID-19, there was a perception that funding streams and referral systems may hinder provision. The lack of a tariff for post-ICU follow-up may cause unwarranted variations, which ICU staff believe could be addressed through a ‘reablement after critical care pathway’ similar to that in place for cardiac and stroke rehabilitation.

Again, before the pandemic, communication between primary and secondary care was sometimes poor, and care was hampered by a lack of clarity about responsibilities for meeting various post-ICU patients’ needs. GPs expressed a need for specific information about recovery from critical illness, collated by a single, authoritative professional group to avoid “*guideline fatigue*” (GP1004, London). All of these existing constraints were believed to have been magnified by the COVID-19 pandemic.

### Strengths and limitations of the study

To the best of our knowledge, this is the first study exploring NHS staff views on follow-up services post-ICU and plans to support patients recovering from severe COVID-19.

We cannot guarantee that our sample is representative of the UK. Responses to our ICU survey were spread across the country, cover different unit sizes, increases in capacity and sizes of NHS Trusts, and are similar to those reported by Connolly, et al. ^14^ GP responses to the survey and interviews was low but they were spread geographically, and all agreed on the challenges of organising care for patients discharged after an ICU stay. Our GP interview findings were consistent with each other, and similar to those with larger samples conducted before the pandemic.^15^

### Meaning of the study: possible explanations and implications for clinicians and policymakers

A number of issues raised in this study are long standing: inadequate discharge summaries, lack of clarity of responsibility for post-acute patient care, fragmented and delayed communication and limited knowledge regarding the support needs of post-ICU patients. During the pandemic, there has been RCGP training about the main post-ICU sequelae, and potential treatments.^16 17^ Problems in continuity of care, however, may need a joint approach to improve local organisation of care.

Community rehabilitation services were described as “patchy”, with long waiting lists; an issue recognised by NHS England.^18^ Recent initiatives to improve provision were welcomed, but some interviewees questioned whether the criteria for determining community rehabilitation needs were fit for purpose for younger, fitter populations, and whether community rehabilitation services could change provision without extra funding to enhance infrastructure. Community mental health services were particularly recognised as overstretched with long waiting lists that prioritise patients at high risk of harming themselves or others.^19^ Murray, et al. ^20^ suggest a model such as the Nightingale hospital, but for rehabilitation, during and after the pandemic.

Commissioning and funding streams seem to be a major issue, as follow-up is recommended but not directly funded, unlike the pathways for cardiac and stroke rehabilitation, which were suggested by interviewees as models for post-ICU care. The evidence base for post-ICU follow-up is however partial and would benefit from further research. ^21 22^

Our interviewees suggested that most of the long-term consequences faced by COVID-19 patients are similar to those faced by others experiencing ICU. Knowledge about the sequelae of COVID-19 is at an early stage, research on longer-term consequences of COVID-19, such as the Post-hospitalisation COVID-19 study (PHOSP-COVID) following more than 10,000 patients for more than 12 months, will shed light on which sequelae relate to being critically ill more generally and which are specific to COVID-19. This should complement what is already known about PICS and effective treatment models.^23^

One interviewee (Bruce J, personal communication) reported that patients who received CPAP reported worse mental health than patients ventilated invasively. According to the Intensive Care National Audit and Research Centre (ICNARC) report to the 9th of October, 44% of COVID-19 patients in critical care settings were not mechanically ventilated during the first 24 hours,^24^ therefore a high proportion of patients are awake and aware of their surroundings. This is significant because, depending on the criteria for prioritisation, this group may not qualify for long-term follow-up, and consequently, might suffer from significant mental health symptoms without receiving formal support. Given the widespread management of COVID-19 patients with CPAP and high flow nasal cannulas, this cohort may need at least as much follow-up as those more invasively ventilated.

### Unanswered questions and future research

Follow-up services vary greatly but the extent to which variations in provision are linked to differences in long-term outcomes is not clear. Identifying models of care which yield the best outcomes in the most efficient way could help develop the evidence base for reducing unwarranted variation in the future. The potential effect on mental health of receiving CPAP while in intensive care also merits further research.

Patients who have had an ICU stay might show impairments even five years after discharge. Currently, appropriate length of follow-up is unclear, as is the point at which care should be continued in primary and community care settings only. Current NICE guidance^25^ addresses the early stage of follow-up, but not longer-term support.

The large cohort of younger than average ICU patients provides an opportunity to assess these services and ensure they meet the needs of those recovering from COVID-19 and other future patients discharged from intensive care.

## Data Availability

Data from the interviews and survey responses are available upon reasonable request

## Author contributions

This study was designed and conceived by ACA, LJ and KB. VD conducted the survey data analysis. ACA and LJ conducted the interviews and their analysis. ACA wrote the first draft of this manuscript. LJ and KB made comments on all the versions of this manuscript. All authors have read and agreed the final version.

## Transparency statement

ACA affirms that this manuscript is an honest, accurate, and transparent account of the study being reported; that no important aspects of the study have been omitted; and that any discrepancies from the study as planned (and, if relevant, registered) have been explained.

## Disclaimer

The views expressed in this article are those of the authors and not necessarily those of the NIHR or the Department of Health and Social Care.

## Data availability statement

Data from the interviews and survey responses are available upon reasonable request.

## Dissemination to participants and related patient and public communities

The findings of the wider programme of research were shared with all our participants to check our interpretations captured their views. All the participants that replied confirmed our interpretations were accurate.

## Ethics approval

This project was reviewed and approved by the Department of Health Ethics Committee at the University of York.

## Acknowledgments

We would like to acknowledge the contribution and advice of the Faculty of Intensive Care Medicine, the Intensive Care Society, the Royal College of General Practitioners, Dr Philip Anthill, Dr Shanthi Anthill, Prof Yvonne Birks, Dr Bronwen Connolly, Dr Tom Lawton, Prof Hugh Montgomery, Prof Rupert Pearse, Prof Zudin Puthucheary, Prof Trevor Sheldon, and Prof Najma Siddiqi.

## References

1. Rengel KF, Hayhurst CJ, Pandharipande PP, et al. Long-term Cognitive and Functional Impairments After Critical Illness. Anesthesia & Analgesia 2019;128(4):772–80. doi: 10.1213/ane.0000000000004066

2. Desai SV, Law TJ, Needham DM. Long-term complications of critical care. Critical Care Medicine 2011;39(2):371–79. doi: 10.1097/CCM.0b013e3181fd66e5

3. Needham DM, Davidson J, Cohen H, et al. Improving long-term outcomes after discharge from intensive care unit: Report from a stakeholders’ conference*. Critical Care Medicine 2012;40(2):502–09. doi: 10.1097/CCM.0b013e318232da75

4. Herridge MS, Tansey CM, Matté A, et al. Functional Disability 5 Years after Acute Respiratory Distress Syndrome. New England Journal of Medicine 2011;364(14):1293–304. doi: 10.1056/NEJMoa1011802

5. Van Aerde N, Meersseman P, Debaveye Y, et al. Five-year impact of ICU-acquired neuromuscular complications: a prospective, observational study. Intensive Care Med 2020;46(6):1184–93. doi: 10.1007/s00134-020-05927-5

6. Pfoh ER, Wozniak AW, Colantuoni E, et al. Physical declines occurring after hospital discharge in ARDS survivors: a 5-year longitudinal study. Intensive Care Med 2016;42(10):1557–66. doi: 10.1007/s00134-016-4530-1

7. Bienvenu OJ, Friedman LA, Colantuoni E, et al. Psychiatric symptoms after acute respiratory distress syndrome: a 5-year longitudinal study. Intensive Care Med 2018;44(1):38–47. doi: 10.1007/s00134-017-5009-4

8. Hodgson CL, Udy AA, Bailey M, et al. The impact of disability in survivors of critical illness. Intensive Care Med 2017;43(7):992–1001. doi: 10.1007/s00134-017-4830-0

9. McPeake J, Mikkelsen ME, Quasim T, et al. Return to Employment after Critical Illness and Its Association with Psychosocial Outcomes. A Systematic Review and Meta-Analysis. Annals of the American Thoracic Society 2019;16(10):1304–11. doi: 10.1513/AnnalsATS.201903-248OC

10. Hatch R, Young D, Barber V, et al. Anxiety, Depression and Post Traumatic Stress Disorder after critical illness: a UK-wide prospective cohort study. Crit Care 2018;22(1):310–10. doi: 10.1186/s13054-018-2223-6

11. FICM. Recovery and rehabilitation for patients following the Pandemic: Faculty of Intensive Care Medicine; 2020 [Available from: https://www.ficm.ac.uk/sites/default/files/ficm_rehab_provisional_guidance.pdf accessed 20 December 2020.

12. Wherton J, Greenhalgh T. Evaluation of the Attend Anywhere / Near Me video consulting service in Scotland, 2019-20. In: Government TS, ed. Social Research Series, 2020.

13. George PM, Barratt SL, Condliffe R, et al. Respiratory follow-up of patients with COVID-19 pneumonia. Thorax 2020;75(11):1009–16. doi: 10.1136/thoraxjnl-2020-215314

14. Connolly B, Douiri A, Steier J, et al. A UK survey of rehabilitation following critical illness: implementation of NICE Clinical Guidance 83 (CG83) following hospital discharge. BMJ Open 2014;4(5):e004963. doi: 10.1136/bmjopen-2014-004963

15. Bench S, Cornish J, Xyrichis A. Intensive care discharge summaries for general practice staff: a focus group study. British Journal of General Practice 2016;66(653):e904–e12. doi: 10.3399/bjgp16X688045

16. RCGP. COVID-19: The patient journey from rehabilitation to recovery: Royal College of General Practitioners; 2020 [Available from: https://elearning.rcgp.org.uk/mod/page/view.php?id=10822 accessed 26 Nov 2020.

17. RCGP. COVID-19: The Patient Journey Through Intensive Care: Royal College of General Practitioners; 2020 [Available from: https://elearning.rcgp.org.uk/mod/page/view.php?id=10801 accessed 26 Nov 2020.

18. NHS RightCare. RightCare: Community Rehabilitation Toolkit, 2020.

19. Naylor C, Bell A, Baird B, et al. Mental health and primary care networks: Understanding the opportunities: The King’s Fund, 2020.

20. Murray A, Gerada C, Morris J. We need a Nightingale model for rehab after covid-19: HSJ; 2020 [Available from: https://www.hsj.co.uk/commissioning/we-need-a-nightingale-model-for-rehab-after-covid-19-/7027335.article accessed 9th October 2020.

21. Geense WW, van den Boogaard M, van der Hoeven JG, et al. Nonpharmacologic Interventions to Prevent or Mitigate Adverse Long-Term Outcomes Among ICU Survivors: A Systematic Review and Meta-Analysis*. Critical Care Medicine 2019;47(11):1607–18. doi: 10.1097/ccm.0000000000003974

22. McIlroy PA, King RS, Garrouste-Orgeas M, et al. The Effect of ICU Diaries on Psychological Outcomes and Quality of Life of Survivors of Critical Illness and Their Relatives: A Systematic Review and Meta-Analysis. Critical Care Medicine 2019;47(2):273–79. doi: 10.1097/ccm.0000000000003547

23. Waldmann CS. Intensive after care after intensive care. Current Anaesthesia & Critical Care 1998;9(3):134-39. doi: https://doi.org/10.1016/S0953-7112(98)80007-0

24. ICNARC. ICNARC report on COVID-19 in critical care: England, Wales and Northern Ireland, 9 October 2020. London, 2020.

25. NICE. Rehabilitation after Critical Illness. NICE Clinical Guideline 83. London, UK: National Institute for Health and Care Excellence, 2009.

